# Barriers and facilitators to implementing social prescribing in child and adolescent mental health services: a qualitative analysis using the Consolidated Framework for Implementation Research

**DOI:** 10.64898/2026.04.29.26349161

**Authors:** Alexandra Bradbury, Emeline Han, Alexandra Burton, Daniel Hayes, Joely Wright, Het Stuttard, Joanna Page, Lou Sticpewich, Daisy Fancourt

## Abstract

**Introduction:** Interventions are urgently needed for young people waiting for Child and Adolescent Mental Health Services (CAMHS) in England. Long waits can worsen mental health, increase distress for young people and families, and place additional pressure on already stretched services. Social prescribing, a referral system for connecting individuals with resources in the community via one-to-one support from a link worker, has not been routinely implemented or evaluated for young people on CAMHS waitlists. It remains unclear whether, and under what conditions, social prescribing can be implemented successfully within CAMHS.

**Methods:** We conducted semi-structured interviews with 23 staff and link workers involved in implementing social prescribing at 11 CAMHS sites across England as part of a large hybrid type II implementation-effectiveness study. We used a framework analysis approach, deriving the coding from the updated Consolidated Framework for Implementation Research (CFIR).

**Results:** Barriers and facilitators mapped to 12 CFIR constructs, generating 26 themes. Key areas included: the challenges of implementing a non-medical intervention in a clinical environment; the advantage of social prescribing compared to little waitlist support; the need for flexibility in mode, duration, and frequency of sessions; the importance of community assets, funding and external partnerships for delivery; and the capacity, skills, and professional experience of link workers and staff. Barriers within CAMHS related to limited resources and partial understanding of the intervention, as well as difficulties in integrating link workers and providing supervision. Successful implementation depended on tailoring the intervention to the needs and preferences of young people and parents. Alternative social prescribing pathways were proposed, with schools being recommended as a promising setting for preventive delivery or post-treatment transitions for young people.

**Conclusion:** Youth social prescribing for young people on CAMHS waitlists is feasible but requires careful implementation. Successful delivery depends on the capacity of link workers and supportive organisational structures in CAMHS. Alternative pathways, including delivery outside the waitlist through schools may facilitate its implementation and impact.

## Introduction

The state of youth mental health in the UK is at its worst in decades. In 2023, NHS England estimated that 1 in 5 young people between the ages of 8 and 16 had a probable mental health disorder (1). Similarly, a 2025 study showed a 10% increase in common mental disorders between 2009-2019 among young adults aged 16-39, with the steepest increases occurring among adolescents aged 16-19 (2). Several factors may be linked with this rise, including the rising cost of living (3), the COVID-19 pandemic (4) (5) (6), increasing rates of adolescent loneliness across countries including the UK (7), and a lack of specialist mental health services in areas of greatest need (8). The growing mental health burden in this age group has been strongly felt by the UK’s NHS Child and Adolescent Mental Health Services (CAMHS), where waitlists for treatment can be as long as 2 years (9). Waiting can exacerbate a young person’s already poor mental health (10). Whilst there are government efforts to expand mental health support for young people outside of CAMHS (11), interventions are needed to support young people once they have reached CAMHS and been told to wait.

“Social prescribing” is a care initiative aimed at connecting patients to community resources via a “link worker.” It focuses around “what matters to you” conversations and connects individuals with non-medical support (e.g., physical, social, and cultural activities). Social prescribing has shown to be effective in adult care, particularly for supporting mental health, managing long-term conditions, and reducing social isolation (12). Link workers play a key role in this process by helping people to identify issues that affect their health and helping them to access resources to meet these needs (13). While social prescribing is well established in adult services, youth-focused models are newer and have been adopted more slowly and inconsistently (11). A recent scoping review demonstrated that there is small but growing evidence of positive outcomes for young people, including for mental health, wellbeing and loneliness (14). Social prescribing can help young people with mental health difficulties to feel supported, discuss their needs, and develop agency over their condition (15), potentially making it highly suitable for those on CAMHS waitlists. However, social prescribing involving one-to-one support from a link worker has not previously been examined in CAMHS.

This study is part of Wellbeing While Waiting (16), a hybrid type II implementation-effectiveness study (16). Clinical outcomes, (reported elsewhere (17)) show that social prescribing was linked with higher resilience and a reduction in difficulties among young people on waitlists, but not with any change in anxiety, stress or depressive symptoms. Given the complexity of social prescribing as a personalised care approach, and the difficulty of examining it in real world environments, broader contextual factors may have influenced its effectiveness. Robust examinations of implementation factors, in combination with impact studies, are needed to understand when, how and in what contexts it can be optimally delivered to improve young people’s mental health outcomes.

The Consolidated Framework for Implementation Research (CFIR) is one of the most well-known determinant frameworks for examining contextual factors that influence implementation (18). It is used to inform strategies to overcome barriers and support future implementation, particularly in health care settings, and it has previously been used in CAMHS (15) (19) (20). The purpose of this qualitative investigation is to use CFIR to explore staff and link worker perspectives regarding the barriers and facilitators that helped or hindered successful implementation of social prescribing for young people on CAMHS waitlists in the Wellbeing While Waiting study.

## Methods

### Study design and sampling

We conducted a multi-site qualitative process evaluation of implementation of the Social Prescribing Youth Network (SPYN) pathway in CAMHS for young people awaiting psychological treatment. The study involved 10 CAMHS sites across England. Full trial procedures are detailed in the Wellbeing While Waiting published study protocol (16). This paper reports on qualitative findings from interviews with CAMHS staff, staff from partner organisations such as public authorities or voluntary, community and social enterprises (VCSE), and link workers regarding the implementation of the pathway.

Participants were eligible if they were aged 18 or over, worked as a CAMHS or NHS staff member, staff member of a partner organisations, or link worker at one of the participating sites, and had capacity to give informed consent. Purposive sampling was used to ensure variation in socio-demographic characteristics, geographical location, and professional role. Selected participants were invited to an interview via email. Written informed consent was obtained from those who indicated they were interested. Participants were either offered a video call interview via Microsoft Teams or a phone call at a time that suited them. Link workers were either employed within CAMHS or seconded from partner organisations. CAMHS staff involved in the intervention had varied roles including oversight, research support, link worker supervision, and assessing and referring young people for social prescribing.

### Intervention

Social prescribing was offered to young people on CAMHS waitlists to provide non-clinical support for their wellbeing while they waited for formal treatment from CAMHS clinicians. The intervention involved up to six sessions with a link worker aimed at helping young people identify their values, interests, and goals, and discussing how these could be achieved through participation in activities in the community. Sessions varied in duration and frequency depending on the young person’s needs and preferences. Link workers helped young people access activities and resources by liaising with community organisations and/or accompanying young people to activities.

### Data collection

We used CFIR 1.0 to structure topic guides as data collection began prior to our analytic phase. We then used CFIR 2.0 for analysis due to its expanded constructs and improved applicability across settings. For the topic guide, questions broadly aligned with the five original CFIR 1.0 domains: intervention characteristics, outer setting, inner setting, characteristics of individuals, and process of implementation. Semi-structured interviews were conducted by ABr, ABu, EH, HS, JP, and JW (see COREQ checklist in Appendix 1 for more details). Interviewers used a topic guide (Appendix 2) to explore participants’ experiences of implementing or delivering social prescribing, including implementation barriers and facilitators, as well as perceived impact of social prescribing. Interviews took place from April 2024 to May 2025 and lasted between 29 to 56 minutes. All interviews were conducted via Microsoft Teams, video-recorded, transcribed verbatim and de-identified. Data was also collected on each participant’s geographical location, age, gender, ethnicity, and professional role. Interviews ceased when the researchers agreed that the data offered sufficient “information power” (21) based on study aim specificity, sample specificity, and thematic richness, to make meaningful comparisons of social prescribing implementation and delivery across different roles and sites.

### Data analysis

Framework analysis was adopted (22), following the stages of data familiarisation, coding, framework identification, framework application, mapping and interpretation. First, transcripts were read by ABr and EH and initial thoughts about the data were noted and discussed. Next, four transcripts that reflected a range of sites, roles, and experiences were independently and inductively coded by ABr and EH. Codes and how they mapped to the CFIR constructs and domains were then discussed and agreed with reference to definitions in the CFIR 2.0 Outcomes Codebook (18). Then, ABr used the CFIR 2.0 framework to code the remaining transcripts, aided by the CFIR user-guide (23). Once coding was complete, barriers and facilitators were organised into themes within each construct and summarised in a table with supporting quotes. Constructs that were not representative of the data were not included.

### Ethics

Ethics approval for the trial was granted by West of Scotland Research Ethics Committee 5 (Ref 22/WS/0184). Informed consent was obtained from all participants.

## Results

Of 75 staff and link workers invited to interviews, 28 consented to take part. Of these, two did not respond, two declined due to limited involvement in implementation, and one did not attend the interview. A total of 23 participants completed interviews, comprising 9 CAMHS staff, 5 partner/external staff, and 9 link workers. (For simplicity, CAMHS staff and partner/external staff are referred to as ‘staff’ throughout.) See Figure 1. For sample demographics see Table 1.

**Figure 1.**
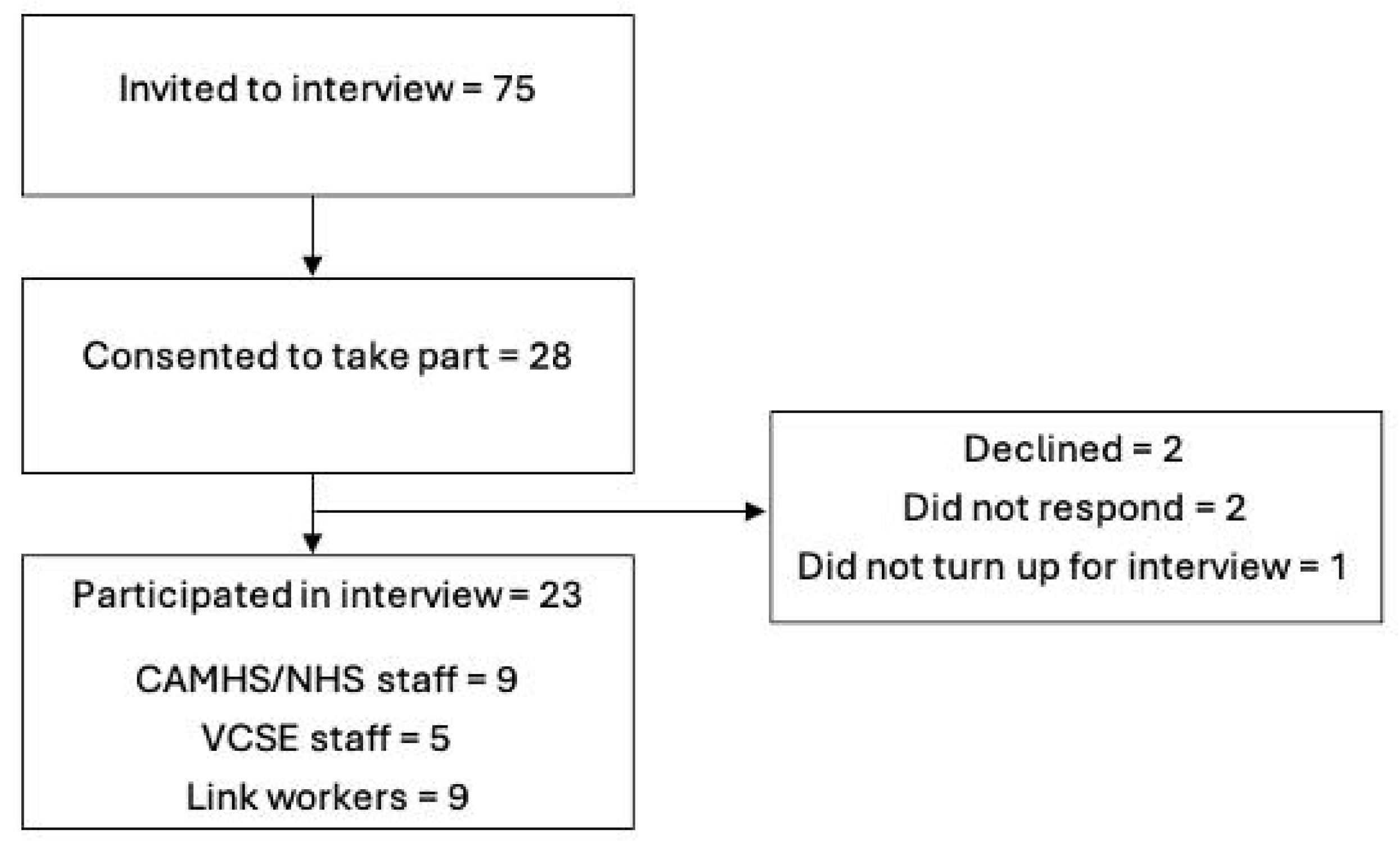
Participant flowchart.

**Table 1:** Characteristics of participants.

| Characteristics | CAMHS/NHS staff (n=9) | External staff (n=5) | Link workers (n=9) | Total |
| --- | --- | --- | --- | --- |
| **Geographical region \*** |  |  |  |  |
| South West | 0 | 1 | 1 | 2 |
| London and South East | 2 | 0 | 3 | 5 |
| North East | 3 | 1 | 2 | 6 |
| North West | 2 | 0 | 4 | 6 |
| East of England | 0 | 1 | 2 | 3 |
| West Midlands | 2 | 2 | 1 | 5 |
| **Role in study \*\*** |  |  |  |  |
| Leadership | 2 | 3 | 0 | 5 |
| Operations / programme management | 3 | 2 | 0 | 5 |
| Research / recruitment | 3 | 0 | 0 | 3 |
| Clinical assessment / referral | 2 | 0 | 0 | 2 |
| Link worker supervision | 2 | 1 | 0 | 3 |
| Social prescribing delivery | 0 | 0 | 9 | 9 |
| **Age range** |  |  |  |  |
| 18-24 | 0 | 0 | 0 | 0 |
| 25-34 | 2 | 0 | 6 | 8 |
| 35-44 | 3 | 1 | 1 | 5 |
| 45-54 | 4 | 3 | 1 | 8 |
| 55-64 | 0 | 1 | 1 | 2 |
| 65+ | 0 | 0 | 0 | 0 |
| **Gender** |  |  |  |  |
| Male | 2 | 2 | 0 | 4 |
| Female | 7 | 3 | 9 | 19 |
| Non-binary/other | 0 | 0 | 0 | 0 |
| **Ethnicity** |  |  |  |  |
| Asian or Asian British | 0 | 0 | 1 | 1 |
| Black, African, Caribbean or Black British | 0 | 0 | 0 | 0 |
| Mixed or Multiple ethnic groups | 0 | 0 | 0 | 0 |
| Other ethnic group | 0 | 0 | 0 | 0 |
| White | 9 | 5 | 8 | 22 |

\*Total location count exceeds number of participants as some link workers operated across multiple geographical regions.
\*\*Total role count exceeds number of participants as some CAMHS/NHS staff had dual roles.

### Domains and constructs

Barriers and facilitators to implementing social prescribing in CAMHS were identified within all five main CFIR domains (Innovation, Outer setting, Inner setting, Individuals, and Implementation Process). Barriers and facilitators mapped to 12 constructs containing 26 themes. To more accurately reflect the context of social prescribing delivery, the Implementation process domain was adjusted so that ‘tailoring strategies’ and ‘engaging recipients’ constructs were merged. See Figure 2. for constructs identified, Table 2 for all themes and barriers and facilitators identified. (See Table S1 in the supplementary material for domain and construct definitions as per the CFIR 2.0 Outcomes Codebook (18).)

**Figure 2.**
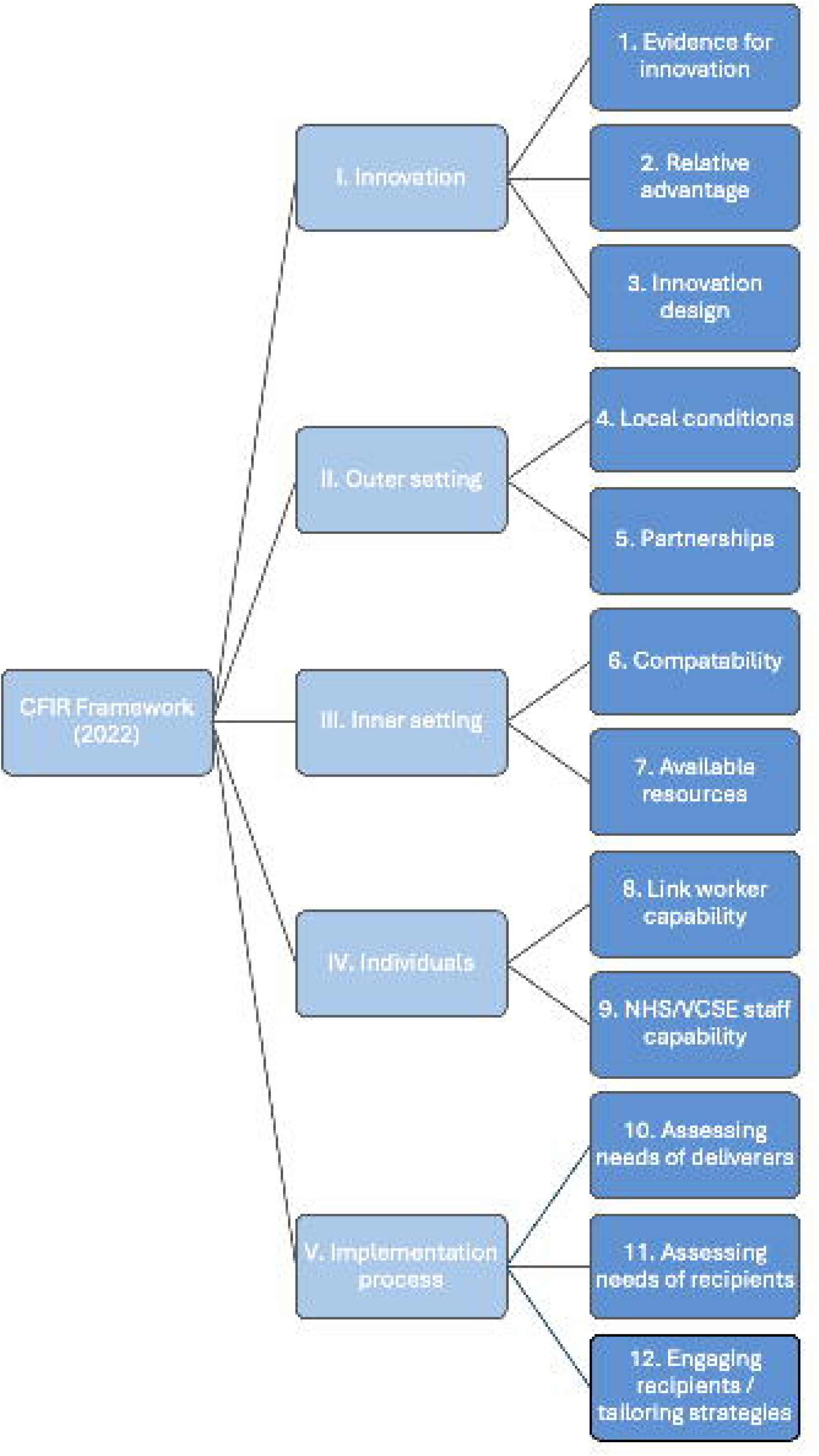
Coding tree showing main constructs identified using the CFIR Framework 2022.

**Table 2:**
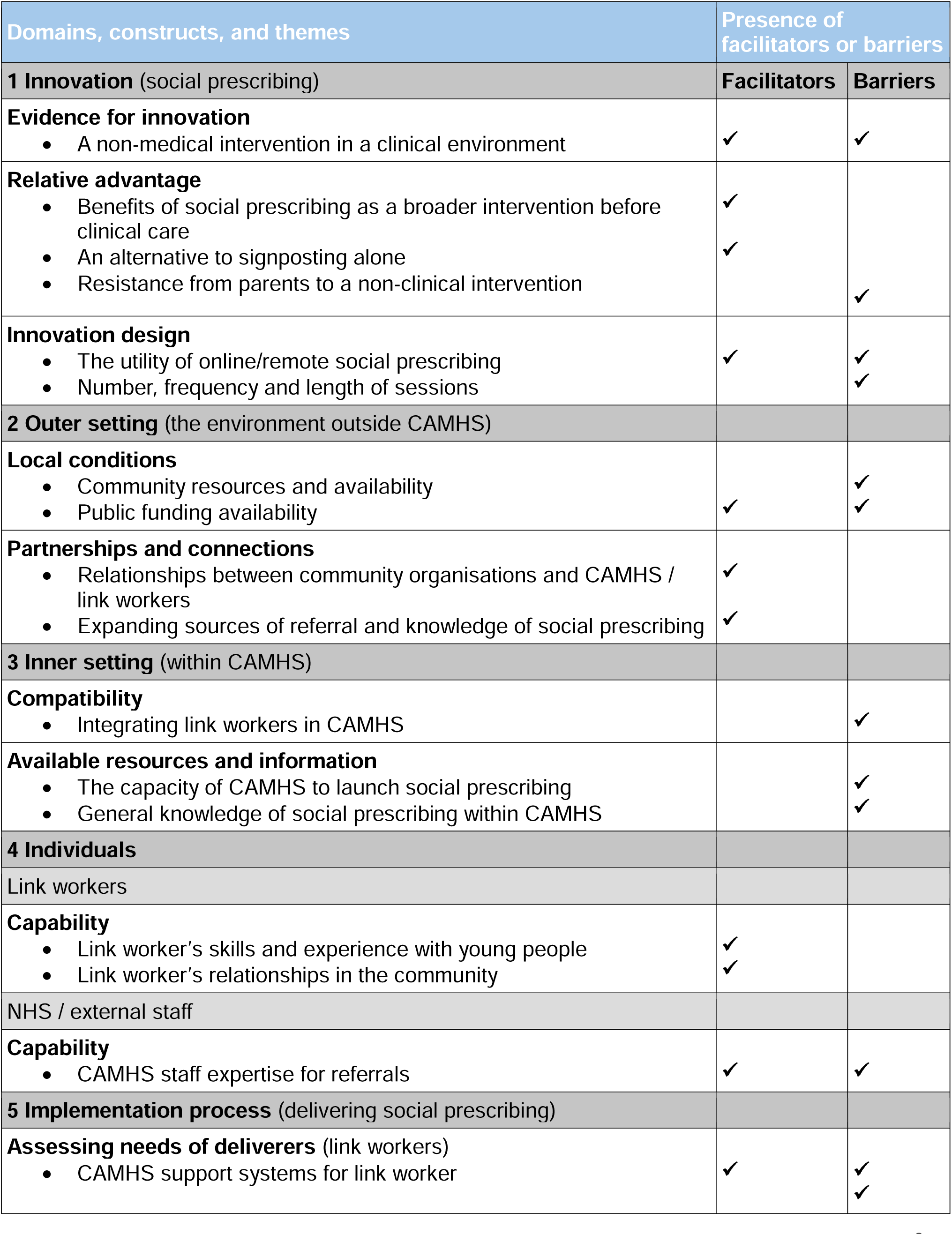
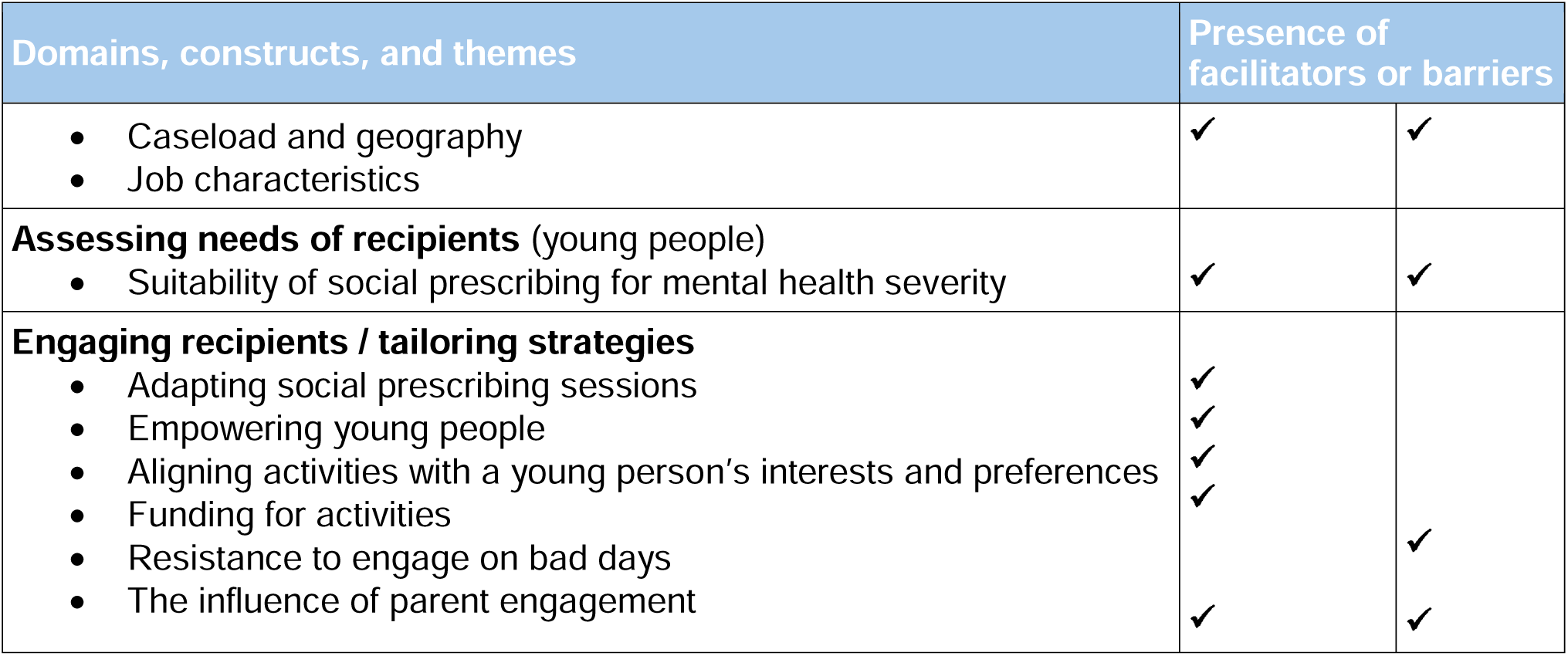
Key implementation factors identified per CFIR construct.

## I. Innovation

### Evidence for innovation

#### A non-medical intervention in a clinical environment

Staff perceived a cultural tension in CAMHS where clinicians expect to practise NICE-aligned and trial-evidenced interventions, a potential barrier for implementing social prescribing given that its evidence base is growing but not yet fully established.

> *NHS services do rely on NICE guidance and things like that. So that I think is a challenge in terms of the research world for any approach, that’s not CBT really, or, or medicine…. we just need the evidence base to come along and strengthen a social I think a social model of, of care. (Staff 23)*

However, for some, the similarity of social prescribing to other evidence-based interventions (e.g., vocational support, mentoring, Occupational Therapy (OT)) legitimised it as an intervention.

> *If you look at the first episode of psychosis service or chronic like long-term illness services, vocational work is a big part… And the vocational worker in essence is a social prescriber. (Staff 24)*

Staff also perceived social prescribing as similar to broader practices such as psychosocial and youth-support work, potentially enabling its implementation.

> *There are other services in that sort of have a similar feel about them. So the youth service offer a life coaching service that I see under that umbrella really. (Staff 5)*

However, this could also mean social prescribing was perceived as non-distinctive, raising concerns about duplication and potentially weakening buy-in.

> *So it’s interesting, is it still social prescribing or does it look more like something else like [occupational therapy], that I’ve been used to?… We don’t invent new… things. We just give them slightly different names, change them slightly. (Staff 23)*

Staff also thought that decision-makers’ desire to have evidence on cost-saving interventions or shorter length of waitlists may be a barrier for future implementation and scaling.

> *From a funding point of view, you’re gonna be looking at how many people did we prevent having to access these services and have we saved money here, there and everywhere. (External staff 11)*

Additionally, broader and longer-term outcomes were highlighted as important so that impact could be measured for a range of health conditions and over years rather than on a project basis.

> *I think there’s something around longitudinal research and outcomes because we know social prescribing is not ‘here, now fixed.’ It’s something I got about giving the young person the building blocks to create a better future… We need to test it out for a range of conditions. Diabetes. Asthma. You know, that kind of things. Clinical labels, because that is speaking the money language. (Staff 13)*

### Relative advantage

#### Benefits of social prescribing as a broader intervention before clinical care

Compared to clinical treatment, social prescribing was perceived by staff as a more holistic option for helping young people with a wide range of issues and giving them support tools while waiting.

> *I think it’s a fantastic service and I think whether it’s engaging young people in social networks and community activity, or whether, as I say, it’s giving them coping strategies and techniques, I think it has a fantastic impact on all young people where they come away with something at the end of that intervention. (External staff 14)*

Social prescribing was perceived by staff and link workers as being especially beneficial for helping young people transition into CAMHS therapy. In particular, if they were struggling to leave the home, speak to new people, or manage anxiety, it was thought that young people might be more receptive to engaging with a clinician after meeting a link worker.

> *In terms of, young people being better prepared, for the clinical work, because they have a better understanding of what it is. They have realistic expectations of what it means. So yeah, we’ve had a lot of success with kind of empowering people to make the most of the clinical work when they do get to that point. (LW 7)*

#### An alternative to signposting alone

Compared to being on a waitlist with limited contact with CAMHS and/or passive signposting, staff and link workers felt that social prescribing was a more proactive option that may help families access community resources while waiting.

> *So I think from that example, the active signposting and the hand holding to cross the threshold is something that works extremely well…Families would receive some signposting but having a live version of that in the form of a care* navigator just changes the probability the families are actually accessing these services. (LW 3)

Staff believed that social prescribing helped parents feel reassured while they had little contact with CAMHS.

> *I know that on CAMHS waiting list, there’s not a lot of contact from CAMHS clinicians and it’s really hard to know like where you are on the waiting list, how long you’re waiting for. And so I think just having a professional in the mix talking to you and keeping in contact is something that makes, I guess it’s just really reassuring often for parents and guardians and families. (LW 19)*

Staff also believed that young people benefitted from the information they gained about resources and activities available in the community.

> *At the very least, I think it points young people in the right direction to other forms of support whilst they’re whilst they’re waiting. (External staff 14)*

#### Resistance from parents to a non-clinical intervention

However, staff commented that sometimes social prescribing was not an appealing offer among parents who placed significant importance on CAMHS therapy, making them less amenable to a link worker’s invitation.

> *I think that we have had some of our families before where they avoid engaging with us sometimes because of not actually understanding what the benefit of it can be, sometimes they quite single-mindedly focused on therapeutic engagement. So, they’re kind of just like we just want to see the therapist. I don’t really want to do any of this. (LW 15)*

### Innovation design

#### The utility of online/remote social prescribing

The option for online delivery of social prescribing was perceived both positively and negatively by stakeholders. Remote social prescribing gave link workers the opportunity to span large geographic areas, and online sessions were sometimes preferred by young people.

> *So a lot of mine had to be done remotely because I was social prescribing for young people in different cities. (LW 18)*
>
> *I had a chat with them and I had a chat with mum. We decided to try doing it online… the young person was just so much more comfortable just being at home and there was less stimuli going on, there was less noise. (LW 17)*

However, online sessions could make it feel more challenging for link workers to engage young people during sessions.

> *They were sitting in, like the family communal room every time we had a session and that meant there was like loads of background noise, there was like younger siblings, like running around and kind of distraction and we have the young person to like, focus on the session and to engage properly. (LW 19)*

Additionally, some link workers felt that not being able to interact with young people in-person and/or accompany them to activities was less impactful.

> *It’s almost a missed opportunity to not be able to be like, ‘Should we just go and get hot chocolate or should we go get a milkshake or walk around this park?’ Even if it’s not getting into an activity it’s just getting them out of the house and having positive experiences. (LW 18)*

Sometimes, online activities that would interest and engage young people were difficult to identify.

> *I found it hard to like come up with activities that felt like the young person was able to take part in it and be really active in it online… So I personally think that having only a remote offer is negative. I don’t think it works. (LW 19)*

Additionally, working remotely could make it more difficult for link workers to contact community organisations from afar.

> *I think the major barriers though would be, so getting people on the phone. Especially if you’re working remotely because it’s easier to go in. If you go in, it’s more likely that people are willing to help you. If you phone up, then people don’t pick up the phone all the time. (LW 17)*

#### Number, frequency and length of sessions

Staff and link workers also commented that the number of sessions (six) and duration of social prescribing sessions (varied by site) was insufficient. Flexibility around the time and the number of sessions was considered very important for having productive experiences with young people.

> *I strongly believe that six sessions isn’t enough. I feel it takes six sessions for lots of kids to learn to trust the adult that they’re working with, let alone be able to talk about things that are really affecting their life. And especially for half an hour sessions, that’s three hours. (LW 17)*

## II. Outer setting

### Local conditions

#### Community resources and availability

The precarity of community organisations was identified as a challenge for implementing social prescribing. Given that many organisations offering youth activities were VCSEs with variable funding sources, their existence could be short-term, making it difficult for link workers to navigate. General operations issues, such as unclear opening hours or lack of response to emails, also posed barriers.

> *Social prescribing depends upon an active and thriving voluntary sector. So it’s all well and good to say we’ve got a team of social prescribers, and we’re gonna refer people out to voluntary sector activities. If that voluntary sector is not adequately resourced and funded you, social prescribers have got nothing to link people in with. (External staff 4)*

#### Public funding availability and responsibility

Relatedly, when public funding was uncertain, this could interfere with social prescribing. For example, given that NHS funding for personalised care budgets (i.e. stipends for young people in this context) has been uncertain, this introduced barriers to launching social prescribing at CAMHS sites.

> *NHS England ended the programme of work, so the personalised care programme of work came to an end. And so I suppose the focus from an NHS perspective dissipated, so that meant we had to work harder to ensure that people remained focused and really kind of understood what it was we were trying to achieve. (External staff 12)*

Additionally, staff expressed concern that in the future, social prescribing may suffer from unclear ownership in terms of who would be responsible for funding and implementing it, whether this be public health authorities or other organisations.

> *Sometimes it’s not owned -- children and young people social programmes -- is not owned by any particular… So I think about adult social prescribing, it’s almost embedded within most primary care networks, whereas children and young people isn’t a standard I don’t find. So for me, I’d like it to see whether it’s primary care networks, whether it’s local councils, whether it is, you know, mental health trusts. (External staff 14)*

### Partnerships and connections

#### Relationships between community organisations and CAMHS / link workers

Given that social prescribing was offered in the outer setting by link workers, the extent that CAMHS relied on partnerships and connections to influence implementation was apparent in the data. Staff reported several facilitators including having productive pre-existing relationships with external VCSE mental health organisations.

> *We’ve worked collaboratively with [organisation] for 20 years… So I think in terms of CAMHS, you know the thing that helped really was this was a pre-existing and long-standing relationship. (External staff 4)*

This allowed them to collaborate more easily, particularly if the VCSE was able to overcome bureaucratic NHS requirements and if both organisations had set up services together before.

> *It was good to work with [VCSE]. They have been marvellous. I don’t know that they always found it very easy to work with us because I was always stuck with bureaucracy or stuff and that kind of things, so that felt quite difficult to working with the inner workings of an NHS Trust has been, and always is. (Staff 23)*

These pre-existing collaborations also enabled CAMHS and their partners to create mutually beneficial networks for information-sharing and support for practitioners.

> *So I’ve built relationship with them over the last, since 19, six years, you know. Creating forums, creating spaces where managers can come together… Any insight I have, I share. (Staff 13)*

Additionally, VCSE partners could be well placed to connect CAMHS and/or link workers with other community organisations to facilitate referrals to social prescribing.

> *The manager of the social prescribing teams in schools (is) absolutely amazing and she is just like this font of knowledge and it was great because I would be able to just e-mail… And she would like, send me like all these links. And that was was just like worth its weight in gold. (LW 9)*

#### Expanding sources of referral and knowledge of social prescribing

CAMHS staff and link workers highlighted the potential benefits of having multiple avenues of referral, allowing other professionals and parents to refer young people to a link worker.

> *I think if the scope is there and there’s the availability for social prescribers to do this outside of the CAMHS service, I definitely think it would be a good thing to do…. (Staff 1)*

Schools, in particular, were frequently mentioned as important partners from where young people could be referred for social prescribing.

> *So I don’t think it should be restricted to people who are waiting for CAMHS treatment… I’d wanna see referrals through mental health support teams. I’d wanna see referrals direct from schools. I’d wanna see referrals from youth projects. (External staff 4)*

However, improving general knowledge and understanding of social prescribing was needed to facilitate implementation in the community, as there was sometimes confusion between social prescribing and other behavioural interventions.

> *I spoke to some people that maybe worked in the NHS or worked in social care that had heard of it, but loads of people were like, no idea what you’re on about, you know. The families, the parents… (Staff 6)*

## III. Inner setting

### Compatibility

#### Integrating link workers in CAMHS

Embedding link workers in CAMHS was considered very important to ensure they had proper supervision, could collaborate on cases with CAMHS staff, and had access to IT and other systems.

> *I think it would work best if link workers were embedded within MDT teams, so I think they should be supported and supervised by clinicians, if this is going to be in CAMHS. (LW 18)*

However, this could be difficult to achieve given NHS regulations and bureaucratic procedures regarding employing new staff and the effort required by a hiring manager to do so.

> *The NHS firewalls are just so hard to get over and getting someone an e-mail address is just really hard. (Staff 1)*

For this reason, some Trusts chose not to employ an in-house link worker but to work with external link workers. However, this could lead to confusion regarding how the link worker would split their time and who they were reporting to.

> *We were generally agreed that it would make better sense for the post to be sat with a voluntary sector and not necessarily within the Trust… [They] probably had a concern and worry that if I was there, how would the needs of [the other place] be reflected in the offer? (External staff 12)*

Future implementation of social prescribing in CAMHS will therefore depend on how clinical and community teams can effectively integrate to ensure strong collaboration, shared goals, and understanding of each other’s value and roles in a young person’s mental healthcare journey.

### Available resources and information

#### The capacity of CAMHS to launch social prescribing

Staff capacity and competing priorities made the implementation of social prescribing within CAMHS difficult. Sometimes, allocating resources to social prescribing was at odds with the needs of other CAMHS projects.

> *It’s really difficult, isn’t it? There’s all the kind of challenges around, you know, where you put resources and we don’t just want to be fighting fires. We do want to be working in an earlier intervention and prevention space, but work predominantly for specialist CAMHS, and, yeah. So it got a little bit political, I think in a way that shouldn’t have. (Staff 23)*

In some case, CAMHS sites simply did not have the funding to employ link workers.

> *They then rejigged the department around and they no longer had the funding, which is why they had to drop out… Throughout the NHS, there is obviously a lack of funding everywhere and we’re, although we’re doing very well as a Trust, recruitment has slowed down like quite a lot in the last couple of months. (Staff 1)*

Allocating current CAMHS staff to a link worker role was sometimes a workaround, but finding staff with the capacity to do so was difficult.

> *And then when she ended up not being able to take on the role, then I had to go to another person. And it’s just finding someone with capacity in CAMHS at the moment was just really difficult. So I think, yeah, one of the main barriers at the moment is sourcing someone internally that would be able to take on that role, would be very difficult. (Staff 1)*

Additionally, social prescribing could be an extra administrative burden for CAMHS staff. For example, making referrals and liaising between link workers and families could be very time-consuming.

> *I think what was a real struggle for us was that it felt quite a big job reviewing the referrals, finding those appropriate referrals and then finding the time within service to ring around all of those families. (External staff 11)*

Once a link worker began their role with CAMHS, their capacity working alone or part-time was often insufficient for the number of young people that could be referred to them.

> *We had two social prescribers and then one left. So I think that that was you know a little bit of it was a lot on one person… Sometimes we maybe had to say no at that point just because again there was only one person and we have to kind of limit that. (Staff 20)*

There was also concern that lack of capacity would make it difficult to launch social prescribing at a larger scale and that proper planning and assessments would need to be undertaken.

> *A full feasibility needs to be done on our end to ensure that this is how this is going to work… This is the whole situation that we’re going to do. Are you actually sure that it’s going to be feasible for us to do it? (Staff 1)*

#### General knowledge of social prescribing within CAMHS

Finally, a general lack of understanding or knowledge of social prescribing among CAMHS staff may have hindered implementation. Staff mentioned difficulties in getting clinicians to understand the potential value of social prescribing, partly due to capacity issues that prevented them from fully absorbing this alternative innovation and taking advantage of it.

> *There’s still a massive job to get clinicians to truly understand the value of social prescribing. And the impact. And there’s a huge challenge in getting them to look up for long enough to understand it. (LW 3)*

## IV. Individuals

### Link workers

#### Capability

##### Link worker’s skills and experience with young people

Link workers’ abilities to identify activities that appealed to the young person and motivate them to participate was crucial to the success of social prescribing.

> *I tried to adapt pretty much all of the activities so in that first session if I found out that a kid liked Star Wars then I would just try and make all of the activities somehow related to Star Wars, so, if I was trying to find out different people in their life that support them, then it would be like, oh, who’s your Obi one Kenobi, or who’s your Darth Vader? (LW 17)*

In addition, the ability to adapt the session and try alternative approaches was also key. Link workers mentioned paying attention to the needs and wants of a young person by observing verbal and non-verbal cues. This included subtle skills like being able to read a young person’s body language to know when to adapt an activity and being as flexible and responsive to a young person as possible.

> *I knew not to pull out my colourful piece of paper with resilience framework written all over it because I just knew she was not going to engage with it. So we just, I just started having those conversations with her and a lot of the answers I was getting were, “I don’t know”, “I don’t care”, “I’m not really bothered”, “I will give it a go, but I can’t promise anything”. So I ended up just changing the conversation I started talking about her, how her nails looked nice, so she warmed up a little bit to me. (LW 3)*

In this process, it was important that link workers were able to develop a trusting and supportive relationship with a young person, which many described as invaluable, regardless of whether the young person also engaged in activities.

> *You know, for a child that has been through a journey of encountering difficulties and having to meet different professionals and then the support comes to an end. Or parents, sometimes they’ll lose trust in the system. So when they get to us, to build these relationships, I feel that this is the most important element. Without trust, there’s no relationship, there’s no engagement, there’s no results. (LW 10)*

Link workers with prior experience in social prescribing, social work, or working with young people described these as attributes that gave them confidence and know-how when engaging with young people. In particular, having experience working with neurodiverse young people was a significant facilitator.

> *One of the things that came up quite a lot was neurodivergency within this project. So that was something that was cited a lot by the young people. And again, the member of staff was, you know, really experienced and was able to point them in the direction of resources and guide them through different things that they could use in terms of coping strategies and techniques. (External staff 14)*

Additionally, link workers had training or experience to recognise safeguarding concerns and escalation of mental health needs beyond their remit, facilitating timely and appropriate involvement of clinical teams.

> *I think you’ve got to be quite a strong person to, to push back and say actually, no, that that’s not social prescribing. And that is too complex and the risks too high. (LW 9)*

Link workers also needed to have a realistic understanding of their role and its limits, and be able to set clear boundaries. Strong communication skills were needed to explain what social prescribing was to parents and young people, increase their understanding of the service and manage their expectations about the scope of support and the goals of sessions.

> *Now you’ve got to tread the fine line between responding to need and not creating dependence… There’s always a kind of an understanding that this is you know, a social prescriber is not a young person’s friend for life… They’re there to help the young person achieve some goals and when those goals have been achieved, the social prescriber needs to step back. (External staff 4)*

##### Link worker’s relationships in the community

Link workers who had already established and maintained prior relationships with community organisations described being well-positioned to introduce young people to activities.

> *Really having that sort of in-depth local knowledge of that community and not just the resources that are available, but almost like the culture of the community that a young person is working in. Like what are those other kind of potential, like socioeconomic things going on in a community that we’re engaging a family or young person with. (External staff 11)*

This was particularly apparent when link workers lived and worked in the same area or had spent considerable time there, which also meant they knew when services had closed or changed.

> *We know people by the first names and I have phone numbers of them that I can just text so the relationships we’ve built are really important because other people keep us up to date about what’s going on in the community as well. (LW 3)*

### CAMHS and partner/external staff

#### CAMHS staff expertise for social prescribing

Given that young people on CAMHS waitlists range in severity and type of mental health condition, staff mentioned that having experience and knowledge to identify appropriate cases to refer to social prescribing was important. This required the ability to understand a young person’s situation and needs, recognise lower-risk cases, and decide who might benefit from a non-clinical intervention.

> *I think, part of that comes from the skill of the people that we’ve got there, but also, the skill of [staff] and the team to identify who’s correct, who’s appropriate. (LW 16)*

It also required the ability to understand the scope of a link worker’s role and whether and how they might be able to help each young person.

> *So there does need to be some consideration about, OK, yeah, complexity and risk. I think there’s also needs to be some thought about, OK, this young person has some goals… this young person wants and wants to get help… and the social prescriber can then sort of take, you know, concrete actions to help though to help make those [goals] become real. (External staff 4)*

However, in reality this was often difficult to do, with CAMHS staff needing to consider multiple issues such as whether one young person might benefit more than another given relative symptoms and needs.

> *But then trying to determine if they were too unwell because it’s like we want to give them the opportunity to have the social prescribing. But then if they’re too unwell, are they going to respond to it and that could take the opportunity away from another child and it was kind of weighing up that was probably the most difficult part. (Staff 1)*

Staff did not always feel they had enough information on a young person to make an informed decision about referring a young person to social prescribing.

> *Say if they’ve they haven’t already had like a full mental health assessment prior to meeting them kind of at this point. You know, just in terms of risk and things that’s sometimes been, you know, a little bit of a, I guess, a difficulty in some sense. (Staff 20)*

Referrals sometimes involved discussions among several staff to determine if a young person might benefit as there was a degree of subjectivity to choosing who to refer.

> *When we’re screening to see people on the wards for research, I’ll be like, well, I don’t think they’re very well, but then someone else might go well, actually, no, I think they’re fine. (Staff 1)*

## V. Implementation process

### Assessing needs of deliverers

CAMHS support systems for link workers

Link workers emphasised the importance of having support and guidance from colleagues and/or a supervisor within CAMHS to discuss and effectively manage cases. Peer support from other link workers or staff, advice from a manager regarding concerning or complex cases, and opportunities to discuss challenges or progress were highly valued by link workers.

> *We had a really good supervisor and group supervisor as well that we could discuss cases with and we also had contacts at the different CAMHS sites that we were working with too. So I was able to work with them, chat with them over the situation and between all of us together we were able to decide that yes, it was appropriate. The young person still wanted to take part, so let’s do it. (LW 17)*

In contrast, when link workers felt unsupported or had no one to raise clinical concerns with, this could cause significant stress and make interactions with parents and young people more difficult. Link workers felt that it was extremely important that a young person’s medical needs be managed by clinical staff and that clear boundaries and monitoring processes be in place.

> *That was really stressful for me because not I wanted, I was happy to do it, but it was leaving me thinking I’m crossing a boundary within my job role here, this is really scary if something happens and I’ve done something wrong, you know that’s a problem because I’m technically not clinical and shouldn’t be doing this. (LW 3)*

This could be made more difficult if the link worker was not embedded in inner workings of CAMHS and did not have adequate information on each of their cases.

> *I think it would be helpful if there was somebody else from CAMHS who is more in touch with the young person in the family…We actually know so little about like what, we’re not embedded enough in the team and in the CAMHS process for to be like the first professional they’re talking with parents and guardians saying like, do you know when sessions are going to like clinically, like the psychological intervention is going to start. (LW 19)*

#### Caseload and geography

Large caseloads and wide geographic areas could overstretch link workers’ capacity in terms of the time they could devote to young people and the numbers they could see.

> *Suddenly you get 3 through at once and you go all right, OK, well, let’s get all of these in and, you know, one’s in [name of town], one’s in [name of town], and one’s in [name of town] and, you know, you’re driving for an hour between each of those spaces, it makes it more complex but it’s a really good, it’s a really important thing to offer, I think. (LW 16)*

Travelling time and caseload challenges were however not always fully appreciated by supervisors or managers.

> *I think in the team of social prescribers I think that having quite a chaotic caseload that was based on lots of different locations was really difficult and a bit stressful at times. I think I was a lot of pressure put on me as a professional to be able to manage that and I think maybe not always an appreciation of how difficult it was to navigate all of that. (LW 19)*

#### Job characteristics

Additionally, if job offers were part-time or short-term, this could be unappealing to link workers looking for full-time or long-term work.

> *We got the agreement for this project and at the same time, all of my social prescribers got made redundant and while one of them said that they’d stay on and do it, they then realised that for one day a week it wasn’t worth their while and they’d got a full-time job… that was the barrier. (LW 16)*

Relatedly, link workers also commented that given the skillset required for their role, salary levels were low and prospects for career progression were unclear.

> *There’s no, clear career progression and so, I think upskilling would be really helpful but also seeing clear career pathways for social prescribers would probably help to keep them. (LW 18)*

Roles that provided opportunities to network and hear from others working in social prescribing were appealing and helpful to link workers.

> *I think you know the buy-in from everybody, project team, CAMHS, you know, workers and our team as well has been great and it’s been lovely to be part of those wider meetings and to hear what’s going on in other spaces, hear the work that you’re doing personally in terms of your, you know, social prescribing and that sort of thing. (LW 16)*

### Assessing needs of recipients

#### Suitability of social prescribing for mental health severity

Staff and link workers expressed concerns that social prescribing *alone* would not be sufficient for young people on the CAMHS waitlist given that their mental health needs were significant enough to warrant clinical support.

> *I don’t think it’ll remove the need for CAMHS if you’re completely if your mental health need is severe enough to actually need proper therapeutic intervention. But I think it’s definitely a really, really important part to run alongside. (Staff 13)*

This was especially true for young people with severe mental health symptoms, who may not be able to cope with the loss of a social prescriber when sessions finished, or who needed ongoing or long-term support.

> *I think to mum, I did just seem a bit naff, you know, ‘why are you talking to my child about fun things when they are suicidal?’ (LW 3)*

As a result, other staff and link workers had stronger views and felt it should not be delivered as a CAMHS waitlist intervention at all.

> *I’m not sure that social prescribing as a waiting list intervention is a good idea. I think there are loads of really positive things that came from it and I think a lot of young people benefited from it. But I think that it being specifically attached to a CAMHS waiting list is not very effective and I think I think it’s just the wrong sort of support at the time. (LW 19)*

Given that social prescribing was better suited for those without significant mental health needs, it was suggested that social prescribing would be better for young people who had not yet reached the CAMHS waitlist.

> *I feel almost like by the time they’ve actually got to CAMHS, they’ve been through so much already. And probably been banging their heads up against the brick wall with GPs and school and stuff. It’s sort of almost like too late. (Staff 6)*

Alternatively, participants suggested that social prescribing could be offered as a step-down approach after CAMHS therapy as a preventative or post-treatment approach.

> *I think social prescribing could be really, really beneficial as, you know, an exit strategy, a step down after treatment… to make sure those treatment gains are maintained and even developed. (External staff 4)*

### Engaging recipients / Tailoring strategies

CAMHS staff and link workers felt that to successfully engage young people in social prescribing, the sessions needed to be tailored in multiple ways.

#### Adapting social prescribing sessions

Link workers who could offer significant flexibility in sessions believed that this was of considerable advantage for engaging young people. This could mean being adaptable with cancellations to encourage families to re-engage despite missed appointments.

> *I would say, I feel we adapt to every single young person or as much as possible. Particularly around being quite accommodating to them not showing up. I feel as much as possible I’ve wanted to give people quite a lot of leeway so that they can not show up repeatedly if that’s what they need, because sometimes they come back. (LW 18)*

Adaptability also involved offering both online and in-person sessions, as well as considering location and transport options for the young person.

> *The way that I got around that is that I had a chat with them and I had a chat with mum. We decided to try doing it online. So we tried it online the next week and the young person was just so much more comfortable just being at home and there was less stimuli going on, there was less noise. (LW 17)*

Link workers also commented on the importance of delivering sessions according to the needs and preferences of the young person. For example, a young person might be willing to have conversations with the link worker but might not be ready to take part in an activity.

> *And I think the majority of my young people weren’t actually at the stage to think about going to an activity or doing it. So the sessions often weren’t really around doing activities in the community and were much more about ‘learning about myself’ or ‘how I’m feeling’. (LW 19)*

Additionally, co-occurring conditions such as autism may require link workers to make additional adaptations to make a young person feel comfortable.

> *I had a session with a young man who has got autism and ADHD… his best friend, his cat, and he likes to sort of mimic what his cat does as well, which very often involves him lying on the top of their American fridge freezer… that is where he wanted to, that’s where he felt most comfortable to engage with me. So, he was up there and I was sat on the kitchen floor. (LW 9)*

When young people were ready to participate in an activity, link workers commented that supporting their first engagement was important, either through accompanying them to the activity, contacting the organisation and requesting they invite the young person and family, or supporting the parent in taking the young person themselves.

> *Sometimes you might have to take them to a service in order for them to engage in it, or you might have to ask the service to reach out to them. You know, so sometimes parents lack confidence in making those phone calls and things. So sometimes if we can say to them, do you want us to ask them to give you a call? Sometimes that can work better. (LW 15)*

#### Empowering young people

Link workers and staff highlighted the importance of giving a young person a sense of “ownership” in sessions. This involved helping young people choose activities and feel like they had direction over what the sessions entailed and what they wanted to talk about so that they had autonomy and more interest in participating.

> *Well, it’s very much kind of client led. So what I say to them in the first appointment is they’re kind of my boss in a way, they love that because it gives them a bit power, and they’re kind of my boss. If they’re interested in anything, it’s my job to go away and then look for that for them and then we’re kind of it just gives them a little bit of ownership, doesn’t it? And the kind of the they’ll buy in a lot better. (LW 8)*

#### Aligning activities with a young person’s interests and preferences

Link workers noticed that young people were more motivated to take part in activities that were personally meaningful or interesting to them.

> *I think I did it with Taylor Swift songs with one as well. So using the things that they’re interested in in order to kind of broaden that chat. First of all, it just makes it easier for them because they can talk about these things with so much information and makes them more comfortable with talking about it because it’s kind of a safe zone. (LW 17)*

Staff and link workers emphasised that the physical location of sessions was important for making a young person feel at ease and able to speak openly. This could include not meeting in public in case the young person was self-conscious about being seen with a link worker.

> *The fact that our social prescriber has been able to go and meet young people in places that are familiar and comfortable for them, that they’re not being asked to come in to, you know, a clinical quite scary space, that they’re getting to choose where they want to meet someone. You know that, I think that goes a long way. (External staff 12)*

#### Funding for activities

Personalised care budgets provided by the NHS for young people to explore an activity was considered vital for some. These monetary stipends allowed them to travel to activities, pay for fees, or buy equipment and was often mentioned as helpful.

> *I think the enabling grants been a big plus, I’ll be honest with you. I think having that little pot of money so that people can try activities, so that they can go out and about. I think it would be a lot more difficult to do if you didn’t kind of have that little bit of financial support. (LW 8)*

#### Resistance to engage on bad days

Sometimes, despite a link worker’s best efforts, the success of a social prescribing session would depend on a young person’s mood. Resistance to engaging could be related to things happening with parents at home or could have no clear cause.

> *So that was really difficult because, she was obviously really angry with Mum. And, and so therefore thought: “I can punish mum by not engaging with [name of interviewee / link worker].” (LW 9)*
>
> *I ended up, like, really panicking and thinking, what am I going to do? You don’t want to do it. I was like, do you want to go to McDonald’s? Do you want to do this? And she said no to everything… It might have just been that she’s just had a bad day. (LW 8)*

#### The influence of parent engagement

A parent’s approach or perception of social prescribing impacted the extent the link worker could engage the young person. Link workers pointed out the benefit of parents who had the interest and time to help a young person investigate and try different community activities.

> *If they have parents who are willing and able to do the homework of, you know, we’ll sit in a session together, this is what I would recommend, let me send you the links… I call them back and they’ve already done it, they don’t need me anymore, they’ve kind of seen value from our first session and just roll with it. (LW 3)*

However, if parents were less involved or supportive of social prescribing, link workers commented on this being a significant challenge for engaging young people. Parents sometimes were unavailable due to work commitments, had their own mental health challenges, or wanted sessions to be adapted in ways that could be at odds with the link worker’s approach.

> *Mum was very much sort of in control. Didn’t want her to have any Teams appointments, wanted me to go over to school after school to see her… But because the young person as well had ADHD, would get distracted very, very easily with everything that was going on. So the sessions, they wasn’t productive… I find sometimes that those young people who don’t engage too well, it can be because the parents are sort of their barrier. (LW 9)*

## Discussion

This multi-site qualitative study identifies key barriers and facilitators to implementing youth social prescribing for young people on CAMHS waitlists, drawing on perspectives from CAMHS staff, VCSE partners and link workers. Our findings reveal fundamental implementation tensions, including balancing youth-centred flexibility with workforce sustainability, navigating organisational resistance to embedding non-clinical care within clinical systems, and resolving uncertainty about where social prescribing belongs in care pathways. Together with findings from the Wellbeing While Waiting evaluation (17), these results help explain why implementation varied across sites and what conditions may enable meaningful support for young people while they wait for care.

### Service design and workforce capacity

Our analysis contributes important evidence to inform the design of youth social prescribing, including the number, length, and mode of sessions. Six sessions were often considered by participants to be too few. While evidence on the optimal social prescribing ‘dosage’ remains inconclusive given intervention variability across contexts (24), data within and beyond this study consistently shows that trusting and supportive relationships with link workers are crucial for effectiveness (25) (26) (27). The importance of allowing sufficient time for relationship-building suggests that session number and duration should be flexible and needs-based rather than fixed.

However, this flexibility can pose challenges for workforce sustainability, with participants reporting a tension between young people’s needs and link workers’ capacities. High work volume for link workers has been reported in other settings (28) (29) (14), and our study reveals that this might be intensified in CAMHS contexts where caseloads span wide geographic areas, young people’s presentations vary in complexity, and partnerships with parents, clinicians, and schools need to be built and maintained. Remote social prescribing, although sometimes helpful for overcoming time and geographic constraints and preferred by some young people, was not considered a replacement for in-person engagement, aligning with adult social prescribing link worker experiences (29).

Factors that enabled link workers to manage their capacity included setting clear boundaries with families and communicating their role and scope of work, indicating that link workers require a strong skillset to strike a careful balance between developing trust and managing expectations. This is in addition to having the ability to respond to individual presentations (body language, mental health symptoms, any co-occurring neurodevelopmental conditions, family circumstances) alongside in-depth local community knowledge (up-to-date information on changing service landscapes and organisational contacts). Link workers and staff in our study emphasised the need for previous experience in relevant roles, aligning with other analyses on link worker competencies (26) (28) (30) (31), while adding that specific experience with neurodivergent young people and established community networks are important facilitators that should be prioritised when hiring youth social prescribers. Our findings also support the need for increased professional recognition, compensation, and career progression commensurate with the rich expertise and sophisticated capabilities expected of link workers (32) (33) (34).

### Cultural acceptance and structural integration

The degree to which social prescribing can be successfully implemented in CAMHS depends on both acceptance within institutional culture and integration within organisational structures. As a non-medical intervention, social prescribing was sometimes met with doubt or resistance in a clinical environment among professionals accustomed to implementing strictly evidence-based interventions such as cognitive behavioural therapy. Yet, staff recognised the benefits of non-medical, tailored support, particularly when framed as analogous to established approaches such as occupational therapy or youth work. This finding echoes other literature documenting that lack of understanding or belief in new initiatives in CAMHS can hinder implementation (35) (36), underscoring the importance of educating clinicians about social prescribing and its value. These cultural tensions also reflect broader debates in youth psychology on the extent to which personalised flexibility can be applied to evidence-based practice for young people (37) (38) (39). Successful youth social prescribing could serve as a strong example of a merged approach, whereby a link worker-clinician partnership for managing cases combines flexible social support with standard clinical treatment.

Beyond cultural acceptance, successful implementation requires structural embedding, including dedicated clinical supervision, IT access, and clear consultation pathways. Our study shows that this was lacking in many cases; link workers often felt removed from the inner processes of CAMHS, lacked access to information needed for their roles, or did not have anyone to consult regarding clinical challenges. Lack of peer support and supervision has been associated with a higher likelihood of link workers leaving their roles (34). In contrast, when link workers were embedded in CAMHS and supported by a clinical manager, they felt able to perform their roles more effectively. Similar observations have been made in studies involving other non-clinical staff in CAMHS; respect, strong collaboration and a treatment climate in which non-clinical staff are valued and have shared goals with clinical staff increases the likelihood of implementation success (40) (35) (41). Our study goes further to show that simply ‘bolting on’ link workers without infrastructure adaptation will undermine implementation.

### Appropriateness and positioning of social prescribing

While staff and link workers perceived social prescribing to have a positive impact on the young people they worked with, they also debated over the appropriateness of social prescribing for young people on CAMHS waitlists. On the one hand, staff considered social prescribing very useful for helping young people to prepare for CAMHS therapy by developing coping strategies and reducing anxiety about clinical appointments. On the other hand, some young people might require more intensive ongoing support and might also struggle with the loss of their link worker once sessions ended. This could help to explain quantitative findings of the wider trial showing improvements in resilience and reduction in total difficulties, but no differences in anxiety, stress, and depressive symptoms between intervention and control groups (17). It also highlights the importance of identifying suitable young people for referral. Staff in this study expressed that referral decisions were not easy to make, and sometimes needed multiple staff to determine if a young person might benefit. The subjectivity of these decisions is a challenge that future research could explore, considering what support can facilitate appropriate social prescribing referrals.

The appropriateness concerns also raise fundamental questions about social prescribing’s optimal position in care pathways. If intended as a low-intensity short-term intervention, it might be better suited for young people reaching the end of the waitlist to transition them into treatment and provide continuity of care. Conversely, given that mental health can deteriorate on waitlists, social prescribing might be better suited as a preventative intervention before families approach CAMHS. Alternatively, it could serve as step-down care after young people leave CAMHS, as suggested by our participants, to maintain treatment gains and facilitate ongoing community connection. Schools were repeatedly mentioned in interviews as important partners for youth social prescribing. While social prescribing involving schools has been evaluated in a preventative care capacity (42), further research could examine CAMHS-school pathways as an after-care model.

Rather than a single model, social prescribing may serve multiple care pathway stages, each with different implications for funding responsibilities and expected outcomes. Participants identified diverse evidence needs across stakeholders: a reduction in demand for CAMHS; proven ability of young people to maintain their wellbeing; improved relationships and confidence; shortened waiting times, and an impact on broader health outcomes beyond mental health. This aligns with other work that has identified core outcome sets for social prescribing (43) (44). However, attempting to serve all purposes simultaneously risks creating unrealistic expectations about what social prescribing can achieve within specific contexts and time frames. Crucially, participants emphasised the importance of measuring young people’s long-term development and successes, tracking young people over time and not just short-term symptom reduction. However, longitudinal evidence requires multi-year funding and evaluation infrastructure, creating tension with current project-based commissioning. Chosen outcome measures and success indicators must therefore align with social prescribing’s specific positioning, intended function, and timeline for impact.

### Strengths and limitations

To the best of our knowledge, this is the first study to examine implementation of social prescribing via a link worker for young people on CAMHS waitlists. It contributes novel evidence on the barriers and facilitators to implementation, and can serve as a guide for youth social prescribing in future healthcare settings. This study employs the updated CFIR Framework (18), which has been adapted to be more applicable to real-world settings. It should be noted that the study and topic guides were developed with the initial version of CFIR, so responses are not perfectly aligned with the new framework. However, we believe the use of the new framework to guide the analysis has resulted in more comprehensive and nuanced results. The hybrid inductive-deductive framework analysis enabled both data-driven theme generation and systematic organisation.

The views gathered in this analysis represent a diverse group of participants including link workers, CAMHS, and VCSE partner organisations who span roles in leadership and management, operations, research, clinical assessment, and delivery. However, of 23 participants, only one was of non-white ethnicity. This highlights a larger problem of link workers not being employed in ethnically diverse areas that have the highest need for social prescribing (1). Additionally, no participants were in the 18-24 age range. This lack of insight from younger people may be a missed opportunity in this study, given that youth peer support in mental health services is increasingly being considered as an impactful way of engaging young people (2). Finally, whilst this study represents the viewpoints of a third of all those invited to participate from the wider Wellbeing While Waiting study, reasons for why individuals chose not to be interviewed are unknown. However, those interviewed contributed extensive diversity in topics and perspectives.

## Conclusion

Implementing youth social prescribing for young people on CAMHS waitlists was shaped by intervention design choices, the integration and support of link workers within CAMHS systems, and the suitability of referrals relative to young people’s level of need. Where supervision, information-sharing, strong understanding and knowledge, and collaborative working were established, stakeholders described greater acceptability and more successful delivery. Future implementation should prioritise workforce support and career pathways, flexible delivery models, and exploration of alternative approaches to delivery, including in schools or through step-down models, all supported by longer-term outcome tracking.

## Supporting information

Table S1. Construct definitions

## Data Availability

As per ethics approval, raw qualitative data cannot be shared due to containing information that might compromise the identity of research participants.

## Declarations

### Ethics approval and consent to participate

Ethics approval for the trial was granted by the NHS Research Ethics Committee (Ref 22/WS/0184). CAMHS staff, partner/external staff, and link workers provided informed consent.

### Consent for publication

All participants provided consent for publication of de-identified data.

### Availability of data and materials

As per ethics approval and consent, raw qualitative data cannot be shared as they contain information that could compromise the identity of research participants.

### Authors’ contributions

DF, DH, and ABu conceived and designed the study. EH, ABr, ABu, JW, HS, JP, and LS conducted interviews. ABr analysed the data with input from other authors (EH, ABu, DF, and DH). ABr drafted the manuscript. All authors reviewed and approved the final version for submission.

## Abbreviations

CAMHS: Child and Adolescent Mental Health Services
CFIR: Consolidated Framework for Implementation Research
VCSE: Voluntary, Community, and Social Enterprise

## Acknowledgements

This paper is independent research supported by the National Institute for Health Research ARC North Thames. The views expressed in this publication are those of the author(s) and not necessarily those of the National Institute for Health Research or the Department of Health and Social Care.

