## Supplementary material for "Barriers and facilitators to implementing social prescribing in child and adolescent mental health services: a qualitative analysis using the Consolidated Framework for Implementation Research": Table S1. Construct definitions

Table S1: Domain and construct definitions as per the CFIR 2.0 Outcomes Codebook

Definitions

**1 Innovation (social prescribing): The “thing” being implemented, e.g., a new clinical treatment, educational program, or city service.**

**Evidence for innovation**: The degree to which the innovation has robust evidence supporting its effectiveness.

**Relative advantage**: The degree to which the innovation is better than other available innovations or current practice.

**Innovation design:** The degree to which the innovation is well designed and packaged, including how it is assembled, bundled, and presented.

**2 Outer setting: The setting in which the Inner Setting exists, e.g., hospital system, school district, state.**

**Local conditions**: The degree to which economic, environmental, political, and/or technological conditions enable the Outer Setting to support implementation and/or delivery of the innovation.

**Partnerships and connections**: The degree to which the Inner Setting is networked with external entities, including referral networks, academic affiliations, and professional organization networks.

**3 Inner setting (CAMHS): The setting in which the innovation is implemented, e.g., hospital, school, city.**

**Compatibility**: The degree to which the innovation fits with workflows, systems, and processes.

**Available resources**: The degree to which resources are available to implement and deliver the innovation. Note: Use this construct to capture themes related to Available Resources that are not included in the subconstructs below.

**4 Individuals: The roles and characteristics of individuals.**

**NHS-VCSE staff:**

- **Mid-level leaders:** Individuals with a moderate level of authority, including leaders supervised by a high-level leader and who supervise others.
- **Implementation facilitators:** Individuals with subject matter expertise who assist, coach, or support implementation.

**Capability**: The degree to which these individuals have interpersonal competence, knowledge, and skills to fulfil Role.

**Link workers**

- **Innovation deliverers:** Individuals who are directly or indirectly delivering the innovation.

**Capability**: The degree to which innovation deliverers have interpersonal competence, knowledge, and skills to fulfil Role.

**5 Implementation process: The activities and strategies used to implement the innovation.**

**Assessing needs of deliverers:** The degree to which individuals collect information about the priorities, preferences, and needs of deliverers to guide implementation and delivery of the innovation.

**Assessing needs of recipients:** The degree to which individuals collect information about the priorities, preferences, and needs of recipients to guide implementation and delivery of the innovation.

**Engaging recipients / tailoring strategies**

The degree to which individuals attract and encourage recipients to serve on the implementation team and/or participate in the innovation

The degree to which individuals choose and operationalize implementation strategies to address barriers, leverage facilitators, and fit context.
